# Comparison of sixteen serological SARS-CoV-2 immunoassays in sixteen clinical laboratories

**DOI:** 10.1101/2020.07.30.20165373

**Authors:** Lene H. Harritshøj, Mikkel Gybel-Brask, Shoaib Afzal, Pia R. Kamstrup, Charlotte S. Jørgensen, Marianne Kragh Thomsen, Linda Hilsted, Lennart Friis-Hansen, Pal B. Szecsi, Lise Pedersen, Lene Nielsen, Cecilie B. Hansen, Peter Garred, Trine-Line Korsholm, Susan Mikkelsen, Kirstine O. Nielsen, Bjarne K. Møller, Anne T. Hansen, Kasper K. Iversen, Pernille B. Nielsen, Rasmus B. Hasselbalch, Kamille Fogh, Jakob B. Norsk, Jonas Henrik Kristensen, Kristian Schønning, Nikolai S. Kirkby, Alex C. Y. Nielsen, Lone H. Landsy, Mette Loftager, Dorte K. Holm, Anna C. Nilsson, Susanne G. Sækmose, Birgitte Grum-Schwensen, Bitten Aagaard, Thøger G. Jensen, Dorte M. Nielsen, Henrik Ullum, Ram B.C. Dessau

## Abstract

Serological SARS-CoV-2 assays are needed to support clinical diagnosis and epidemiological investigations. Recently, assays for the large-volume detection of total antibodies (Ab) and immunoglobulin (Ig) G and M against SARS-CoV-2 antigens have been developed, but there are limited data on the diagnostic accuracy of these assays. This study was organized as a Danish national collaboration and included fifteen commercial and one in-house anti-SARS-CoV-2 assays in sixteen laboratories. Sensitivity was evaluated using 150 serum samples from individuals diagnosed with asymptomatic,mild or moderate nonhospitalized (n=129) or hospitalized (n=31) COVID-19, confirmed by nucleic acid amplification tests, collected 13-73 days from symptom onset. Specificity and cross-reactivity were evaluated in samples collected prior to the SARS-CoV-2 epidemic from > 586 blood donors and patients with autoimmune diseases or CMV or EBV infections. Predefined specificity criteria of ≥ 99% were met by all total-Ab and IgG assays except one (Diasorin/LiaisonXL-IgG 97.2%). The sensitivities in descending order were: Wantai/ELISA total-Ab (96.7%), CUH/NOVO in-house ELISA total-Ab (96.0%), Ortho/Vitros total-Ab (95.3%), YHLO/iFlash-IgG (94.0%), Ortho/Vitros-IgG (93.3%), Siemens/Atellica total-Ab (93.2%), Roche-Elecsys total-Ab (92.7%), Abbott-Architect-IgG (90.0%), Abbott/Alinity-IgG (median 88.0%), Diasorin/LiaisonXL-IgG (84.6%),Siemens/Vista total-Ab (81.0%), Euroimmun/ELISA-IgG (78.0%), and Snibe/Maglumi-IgG (median 78.0%). The IgM results were variable, but one assay (Wantai/ELISA-IgM) had both high sensitivity (82.7%) and specificity (99%). The rate of seropositivity increased with time from symptom onset and symptom severity. In conclusion, predefined sensitivity and specificity acceptance criteria of 90%/99%, respectively, for diagnostic use were met in five of six total-Ab and three of seven IgG assays.

## Introduction

The World Health Organization (WHO) was notified in late December 2019 regarding a cluster of cases of pneumonia in Wuhan City, China. The virus responsible was isolated in the first week of January 2020, and its genome was shared a week later. Phylogenetic analysis showed that it was a novel coronavirus, designated initially as 2019 novel coronavirus (2019-nCoV) and later as severe acute respiratory syndrome coronavirus 2 (SARS-CoV-2). SARS-CoV-2 quickly spread worldwide, and the WHO declared COVID-19 a pandemic on March 11^th^ (1).

In the following months, several hundred assays for detecting SARS-CoV-2 emerged. Different versions of nucleic acid amplification tests (NAATs) for naso-/oro-pharyngeal swabs or washes and lower respiratory tract specimens are the method of choice for detecting COVID-19 (2). However, assays for detecting antibodies that are produced as part of the humoral immune response to SARS-CoV-2 infection have emerged only recently (3). These assays mostly show that one week after the first symptoms, 30% of the patientswith COVID-19 have seroconverted, increasing to 70% after the 2^nd^ week and to above 90% by the 3^rd^ week (4). Accordingly, serological assays measuring either total antibodies (total Ab), immunoglobulin G (IgG) or IgM against antigens of SARS-CoV-2 have been used for supporting a diagnosis of COVID-19, for monitoring the epidemic, and for screening recovered COVID-19 patients to derive plasma for use in convalescent plasma therapy (5). Currently, the numerous serological assays have only been validated on very small numbers of samples and at best have been approved for emergency use with only a few days of evaluation. Several serological assays, especially the lateral flow point-of-care tests, have a suboptimal performance with a low sensitivity and are not recommended for diagnostic use or even for population monitoring (6-8). Recently, several manufacturers of larger instruments have released serological assays useful for mass testing, but few studies have compared these assays directly (9). This comparison is needed for the commutability of the test results and the scientific data. Here, we present a national evaluation of sixteen serological SARS-CoV-2 immunoassays across 16 different laboratories in Denmark.

## Materials and Methods

A case control design was chosen in order to include pre-pandemic samples.

### Case panel samples for determination of clinical sensitivity

The case panel samples tested in all assays (N=150) were obtained from a variety of convalescent patients in the Capital Region of Denmark with a confirmed SARS-CoV-2 NAAT result, that were identified in the Danish national microbiology database (MiBa) from February 2020 to April 2020(10). Individuals were contacted via public secure mail and a total of 639 persons responded. Serum samples of 3.5 ml and 4 ml EDTA samples were obtained from each person in 20 + 2 separate vials during May 3^rd^ to May 11^th^, 2020. For this study serum samples from sample date May 3^rd^ were chosen and send to all participating laboratories. Epidemiologic and clinical data were collected from an electronic self-reported questionnaire completed on the day of blood sampling.

### Archive samples for determination of clinical specificity

Archived plasma samples from pre-COVID-19 blood donations drawn during the influenza seasons of 2017-2018 and 2018-2019 were tested with the SARS-CoV-2 total-antibody (Ab) assays and the IgG and the IgM assays. The number of tested samples (N) were >586 for the total-Ab and IgG assays but only >400 for the IgM assays. Different samples were used across regions, with minor overlap in some cases.

### Archive samples for determination of cross-reactivity

For all assays, cross-reactivity (x-reactivity) was investigated by testing patients with unspecified autoimmune diseases (N=10-131). Due to challenges with available amounts of sample material, 10 samples were pooled and tested across all assays. The nonpooled samples were tested in selected assays. Additionally, for all assays, the archived samples from patients with acute infections of Cytomegalovirus (CMV) or Epstein Bar virus (EBV) or other acute viral diseases were tested (N=10-37), but different samples were used across the assays. All samples were obtained prior to January 2020, before the first COVID-19 case in Denmark.

### Immunoassay platforms

The performance of diagnostic accuracy of commercial immunoassays for the detection of anti-SARS-CoV-2 total Ab, anti-SARS-CoV-2 IgG and anti-SARS-CoV-2 IgM were tested on the appropriate platforms by experienced laboratory technicians following the manufacturers’ protocols with the cut-off values suggested by the manufacturers (Table 1).

**Table 1.**
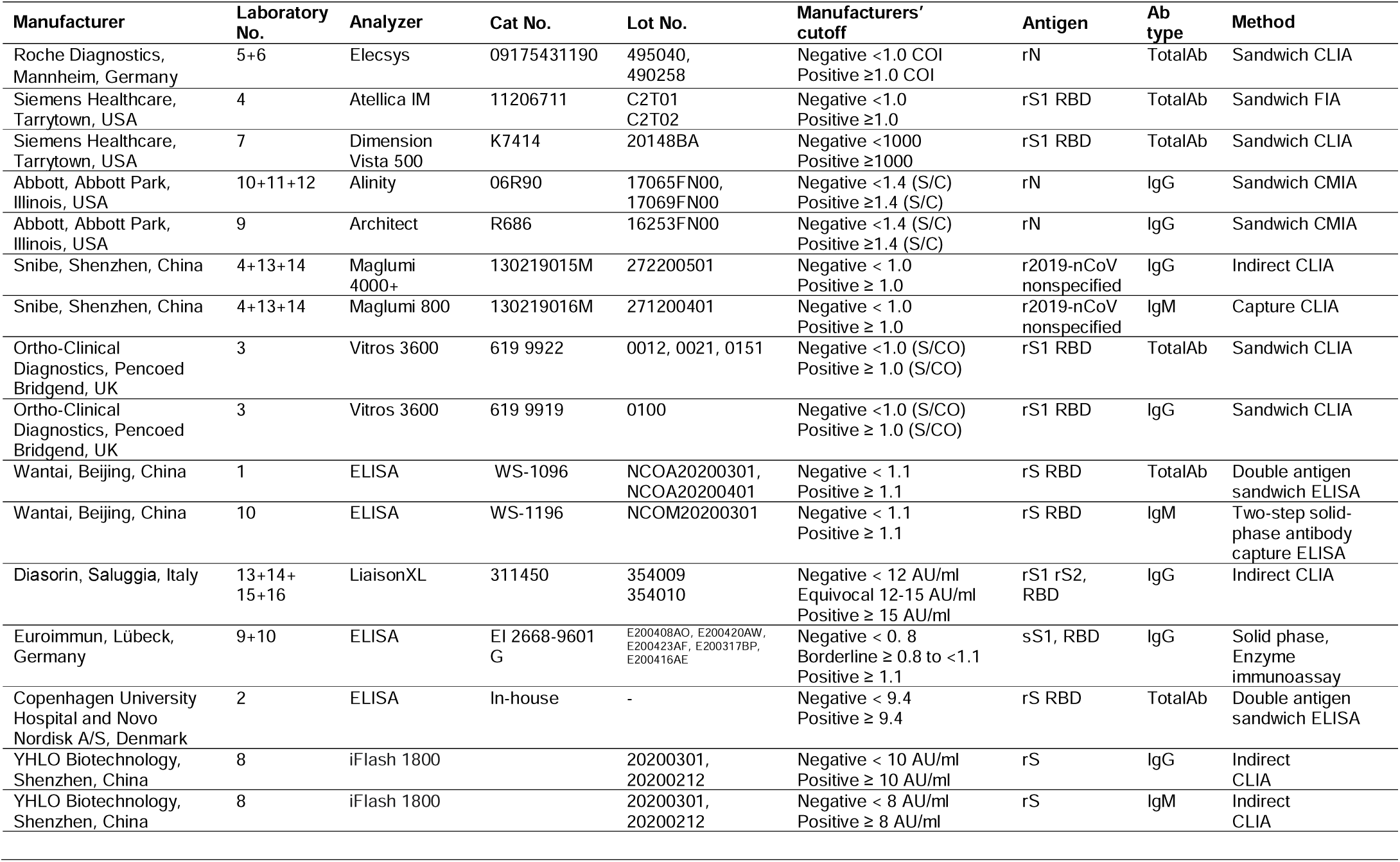
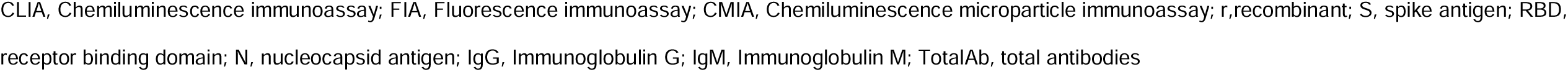

| Manufacturer | Laboratory No. | Analyzer | Cat No. | Lot No. | Manufacturers' cutoff | Antigen | Ab type | Method |
| --- | --- | --- | --- | --- | --- | --- | --- | --- |
| Roche Diagnostics, Mannheim, Germany | 5+6 | Elecsys | 09175431190 | 495040, 490258 | Negative <1.0 COI<br>Positive ≥1.0 COI | rN | TotalAb | Sandwich CLIA |
| Siemens Healthcare, Tarrytown, USA | 4 | Atellica IM | 11206711 | C2T01<br>C2T02 | Negative <1.0<br>Positive ≥1.0 | rS1 RBD | TotalAb | Sandwich FIA |
| Siemens Healthcare, Tarrytown, USA | 7 | Dimension Vista 500 | K7414 | 20148BA | Negative <1000<br>Positive ≥1000 | rS1 RBD | TotalAb | Sandwich CLIA |
| Abbott, Abbott Park, Illinois, USA | 10+11+12 | Alinity | 06R90 | 17065FN00, 17069FN00 | Negative <1.4 (S/C)<br>Positive ≥1.4 (S/C) | rN | IgG | Sandwich CMIA |
| Abbott, Abbott Park, Illinois, USA | 9 | Architect | R686 | 16253FN00 | Negative <1.4 (S/C)<br>Positive ≥1.4 (S/C) | rN | IgG | Sandwich CMIA |
| Snibe, Shenzhen, China | 4+13+14 | Maglumi 4000+ | 130219015M | 272200501 | Negative < 1.0<br>Positive ≥ 1.0 | r2019-nCoV nonspecified | IgG | Indirect CLIA |
| Snibe, Shenzhen, China | 4+13+14 | Maglumi 800 | 130219016M | 271200401 | Negative < 1.0<br>Positive ≥ 1.0 | r2019-nCoV nonspecified | IgM | Capture CLIA |
| Ortho-Clinical Diagnostics, Pencoed Bridgend, UK | 3 | Vitros 3600 | 619 9922 | 0012, 0021, 0151 | Negative <1.0 (S/CO)<br>Positive ≥ 1.0 (S/CO) | rS1 RBD | TotalAb | Sandwich CLIA |
| Ortho-Clinical Diagnostics, Pencoed Bridgend, UK | 3 | Vitros 3600 | 619 9919 | 0100 | Negative <1.0 (S/CO)<br>Positive ≥ 1.0 (S/CO) | rS1 RBD | IgG | Sandwich CLIA |
| Wantai, Beijing, China | 1 | ELISA | WS-1096 | NCOA20200301, NCOA20200401 | Negative < 1.1<br>Positive ≥ 1.1 | rS RBD | TotalAb | Double antigen sandwich ELISA |
| Wantai, Beijing, China | 10 | ELISA | WS-1196 | NCOM20200301 | Negative < 1.1<br>Positive ≥ 1.1 | rS RBD | IgM | Two-step solid-phase antibody capture ELISA |
| Diasorin, Saluggia, Italy | 13+14+15+16 | LiaisonXL | 311450 | 354009<br>354010 | Negative < 12 AU/ml<br>Equivocal 12-15 AU/ml<br>Positive ≥ 15 AU/ml | rS1 rS2, RBD | IgG | Indirect CLIA |
| Euroimmun, Lübeck, Germany | 9+10 | ELISA | EI 2668-9601 G | E200408AO, E200420AW, E200423AF, E200317BP, E200416AE | Negative < 0.8<br>Borderline ≥ 0.8 to <1.1<br>Positive ≥ 1.1 | sS1, RBD | IgG | Solid phase, Enzyme immunoassay |
| Copenhagen University Hospital and Novo Nordisk A/S, Denmark | 2 | ELISA | In-house | - | Negative < 9.4<br>Positive ≥ 9.4 | rS RBD | TotalAb | Double antigen sandwich ELISA |
| YHLO Biotechnology, Shenzhen, China | 8 | iFlash 1800 |  | 20200301, 20200212 | Negative < 10 AU/ml<br>Positive ≥ 10 AU/ml | rS | IgG | Indirect CLIA |
| YHLO Biotechnology, Shenzhen, China | 8 | iFlash 1800 |  | 20200301, 20200212 | Negative < 8 AU/ml<br>Positive ≥ 8 AU/ml | rS | IgM | Indirect CLIA |

CLIA, Chemiluminescence immunoassay; FIA, Fluorescence immunoassay; CMIA, Chemiluminescence microparticle immunoassay; r, recombinant; S, spike antigen; RBD, receptor binding domain; N, nucleocapsid antigen; IgG, Immunoglobulin G; IgM, Immunoglobulin M; TotalAb, total antibodies

### Enzyme-linked immunosorbent assay (ELISA)

The commercial ELISA assays for anti-SARS-CoV-2 total Ab, IgG and IgM detection were performed on open platform analyzers or manually by experienced laboratory technicians according to the manufacturers’ instructions. The Euroimmun SARS-CoV-2 IgG assay was performed on an Analyzer I (Euroimmun AG, Lübeck, Germany), QUANTA Lyser 160 (INOVA Diagnostics, San Diego, CA, USA) or EVOLIS (BioRad, Hercules, CA, USA). The Wantai SARS CoV-2 total-Ab and IgM assays were performed manually and measured using a Tecan Sunrise ELISA reader (Männedorf, Switzerland) at 450 nm with reference at 620 nm.

One in-house ELISA detecting total Ab was tested in this evaluation. Briefly, the *CUH-NOVO SARS-CoV-2 total-Ab* ELISA (a noncommercial assay produced in-house in a collaboration between Copenhagen University Hospital and Novo Nordisk A/S, Denmark) is based on a recombinant receptor-binding domain (RBD) of the SARS-CoV-2 spike protein used both for coating and detection. The assay will be described in detail elsewhere. In this study, the samples were diluted 1:100 in PBS tween-20 in 96-well plates coated with RBD. Total-Ab was detected using HRP-conjugated streptavidin diluted in PBS-T mixed with biotin-labeled RBD. TMB ONE was used as a substrate. The reaction was stopped with 0.3 M H2SO4, and the optical density (OD) of the samples was measured at 450-620 nm. The CUH-NOVO SARS-CoV-2 total Ab ELISA used a semi-automated setup, and the results were based on signal-to-noise (S/N) ratios between the samples of interest and the negative quality control. The cut-off value was calculated based on receiver operating characteristic (ROC) analysis by prioritizing the specificity. Sample results < 9.4 were interpreted as negative. The assay can also be performed in a 384-well format.

Some assays were evaluated in more than one laboratory.

### Statistics

Data handling, graphics and statistics were performed using the R statistical software (v.4.0.2; R Foundation for Statistical Computing, Vienna, Austria (URL https://www.R-project.org/)). The parameters of diagnostic accuracy and the plots were determined using the mada package (meta-analysis of diagnostic accuracy; Philipp Doebler (2019), R package version 0.5.9 (https://CRAN.R-project.org/package=mada)). For calculation of the 95% confidence intervals for the sensitivity and specificity, the default “Wilson” option was chosen. For plotting of bivariate confidence regions in the ROC space, a continuity correction of 0.5 was applied.

### Performance criteria

We defined acceptance criteria for the diagnostic accuracy of the assays depending on immunoglobulin type and intended use. In a low-seroprevalence setting, the specificity of the test is the most important concern and must be high. Irrespective of specific clinical indication for the use of the anti-SARS-CoV-2 total-Ab and anti-SARS-CoV-2 IgG assays, we defined the specificity acceptance criteria to be ≥99%. For the SARS-CoV-2 total-Ab assays, the defined acceptable sensitivity for diagnostic use was ≥92%, with an optimal sensitivity of ≥95%. For the anti-SARS-CoV-2 IgG assays, the defined acceptable sensitivity was ≥90%. For epidemiologic surveys, the acceptable sensitivity was ≥80% for both the anti-SARS-CoV-2 total-Ab and the anti-SARS-CoV-2 IgG assays. No acceptance criteria for the diagnostic accuracy of the anti-SARS-CoV-2 IgM assays were defined, as most of the samples in the case panel were collected > 3 weeks after TSO.

## Results

Of 150 patient samples, epidemiologic and clinical data were received from 149 patients; for the patient characteristics, refer to Table 2. Most of the patients were categorized with clinically mild to moderate symptoms (N=112), and only 31 cases had been admitted to hospital. Time from symptom onset (TSO) was >14 days in 133 case panel patients, while 20 patients had a TSO ≤ 3 weeks and 120 samples were drawn > 3 weeks from the TSO, of which 79 samples (53%) were collected > 6 weeks after symptom onset. Among a total of 7 patients with a TSO ≤14 days, the TSO was 13 or 14 days.

**Table 2.** Case panel patient characteristics

| Characteristic | N (%) |
| --- | --- |
| Sex |  |
| Male | 52 (34.9) |
| Female | 97 (65.1) |
| Age, median (IQR) years | 54 (43-64) |
| Age, range, years | 18-83 |
| Time from symptom onset (TSO), days |  |
| 0-7 | 0 (0.0) |
| >7-14 | 7 (4.7) |
| >14-21 | 13 (8.7) |
| >21-42 | 49 (32.7) |
| >42 | 71 (47.3) |
| NA | 9 (6.0) |
| Time from positive SARS-CoV-2 PCR, days |  |
| 0-7 | 1 (0.7) |
| >7-14 | 15 (10.0) |
| >14-21 | 22 (14.6) |
| >21-42 | 90 (60.0) |
| >42 | 21 (47.3) |
| Symptom degree |  |
| No symptoms | 6 (4.0) |
| Mild (at home, well) | 37 (24.8) |
| Moderate (home, bedridden) | 75 (50.3) |
| Severe (hospitalized) | 2 (1.3) |
| Critical (assisted ventilation) | 29 (19.5) |
| Total | 149 (100) |

N, number; IQR, Interquartile range; NA, not available.

### Data on the detection of anti-SARS-CoV-2 antibodies according to the assay

The results of the tests, i.e., true positives (TP), false negatives (FN), false positives (FP), and true negatives (TN), as well as the calculated sensitivity and specificity of each assay, are presented in Table 3 in descending order of sensitivity, accounting for specificity. The specificities are calculated by combining the data from all sites that validated the same assay. The key to each laboratory in Table 3 is presented in (Table S1). The commercial ELISAs and the Diasorin/LiaisonXL assay have defined gray zone results. In our results and calculations, we defined gray zone results as negative.

**TABLE 3.**
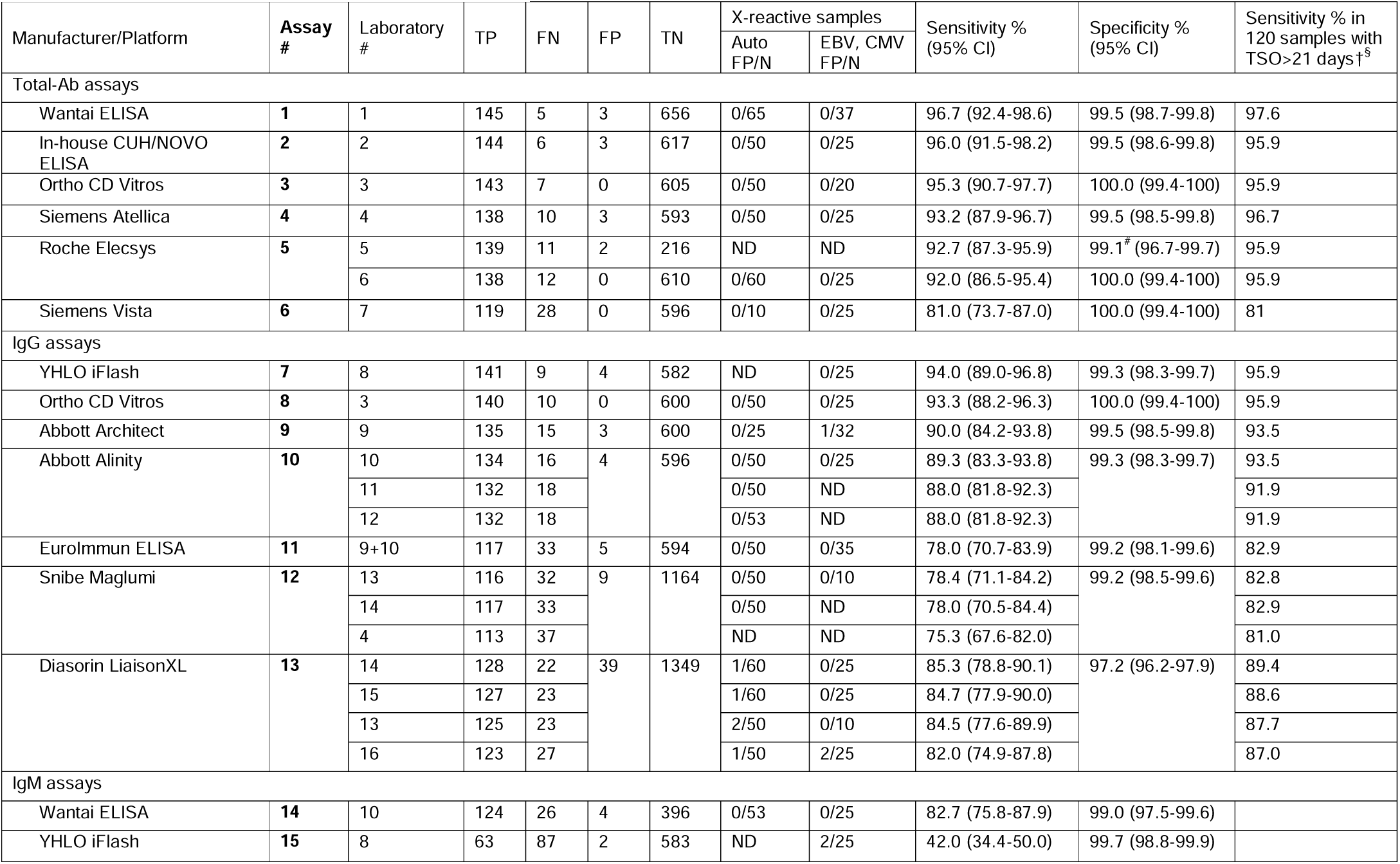
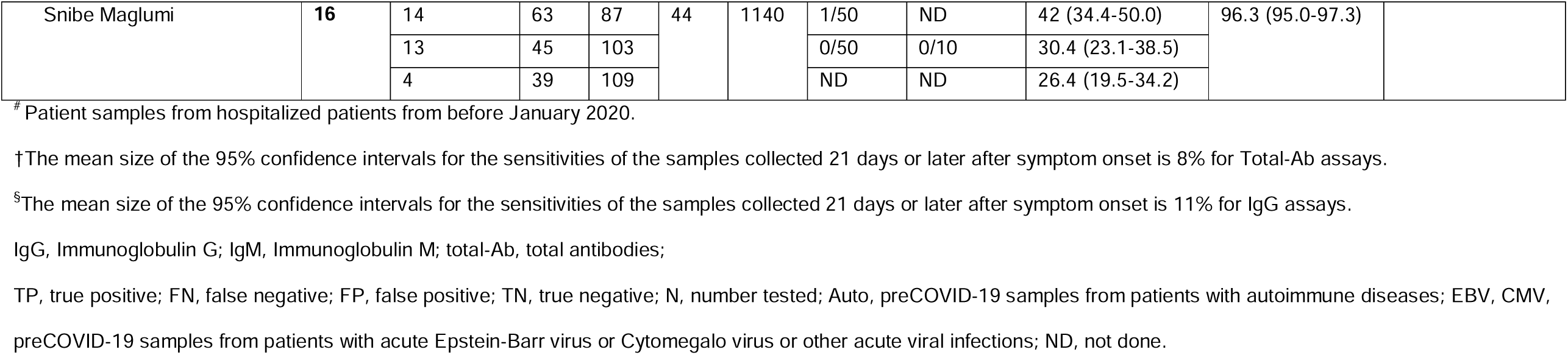
Anti-SARS-CoV-2 antibody assays with results

All total-Ab assays performed with high and acceptable specificities (≥99%). The two total-Ab ELISAs and the Ortho Vitros total-Ab assay performed with an optimal sensitivity (≥95%), while the Siemens Atellica and Roche Elecsys assays performed with acceptable sensitivity for diagnostic use (≥92%). One outlier - the Siemens Vista - performed with an acceptable sensitivity for epidemiologic surveys (≥80%) but not for diagnostic use (≥92%). Of the IgG assays, all but the Diasorin/LiaisonXL IgG assay resulted in acceptable specificities (≥99%). Three assays (the YHLO iFlash IgG, the Ortho CD Vitros IgG and the Abbott Architect IgG assays) resulted in acceptable sensitivities (≥90%). The sensitivity was improved in all total-Ab and IgG assays if the analyses were restricted to samples collected > 3 weeks after symptom onset (Table 3).

Regarding the IgM assays, the performance of the Wantai IgM ELISA demonstrated a much higher sensitivity than the other two IgM assays, with an acceptable specificity of 99.0% and no detected x-reactivity (Fig. 1). The sensitivity of the YHLO/iFlash IgM assay was low, and x-reactivity was detected in 2 of 25 samples from pre-COVID-19 patients with either acute CMV or EBV infections, whereas the Snibe/Maglumi IgM assay performed poorly in specificity and sensitivity (Fig. 1).

**Figure 1.**
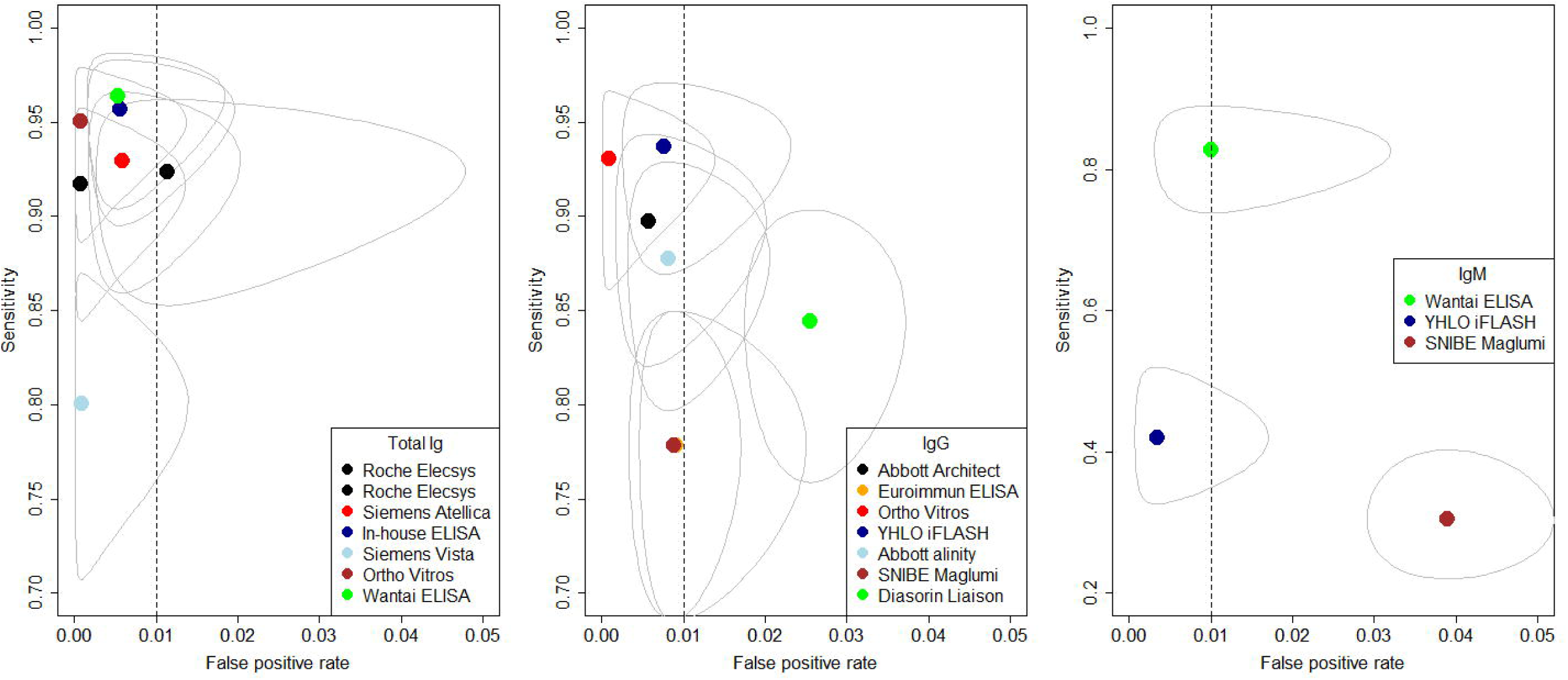
Summary ROC plot of sensitivity and false positive rate with elliptic 95% bivariate confidence regions corresponding to the data in table 3 for assays with total Ig, IgG and IgM respectively. For the IgG assays where data was available from more than one laboratory the median result was chosen for the COVID-19 cases and for the pre-pandemic blood donors the total of all the samples was used, as these were from different persons. The vertical broken line at a false positive rate of 0.01 correspond to a 99% specificity. The y-axis for IgM has a different scale from 20% sensitivity instead of 70%.

### Data on the detection of anti-SARS-CoV-2 antibodies according to TSO and disease severity

The largest variation in true positive samples between the assays for each Ig type category was shown among samples with a TSO ≤21 days (Fig. 2). In addition, increasing rates of seropositive results were found with the severity of symptoms in all assays (Fig. 2). Generally, serological testing had a low sensitivity when tested less than three weeks from symptom onset and in patients who were asymptomatic or had mild disease at home but were not bedridden. If patients have been bedridden or hospitalized, the sensitivity in the convalescent cases by serological testing was >90% for most of the total-Ab or IgG assays included in the study (Fig. 2). Except for one assay (Wantai IgM), the IgM assays showed a very low sensitivity, especially among the patients with mild symptoms. The large difference in sensitivity according to the disease severity was not explained by the difference in time of testing. The two large groups of the nonhospitalized symptomatic cases both had a similar median TSO near 40 days and a range of distribution from symptom onset to blood sampling (results not shown).

**Figure 2.**
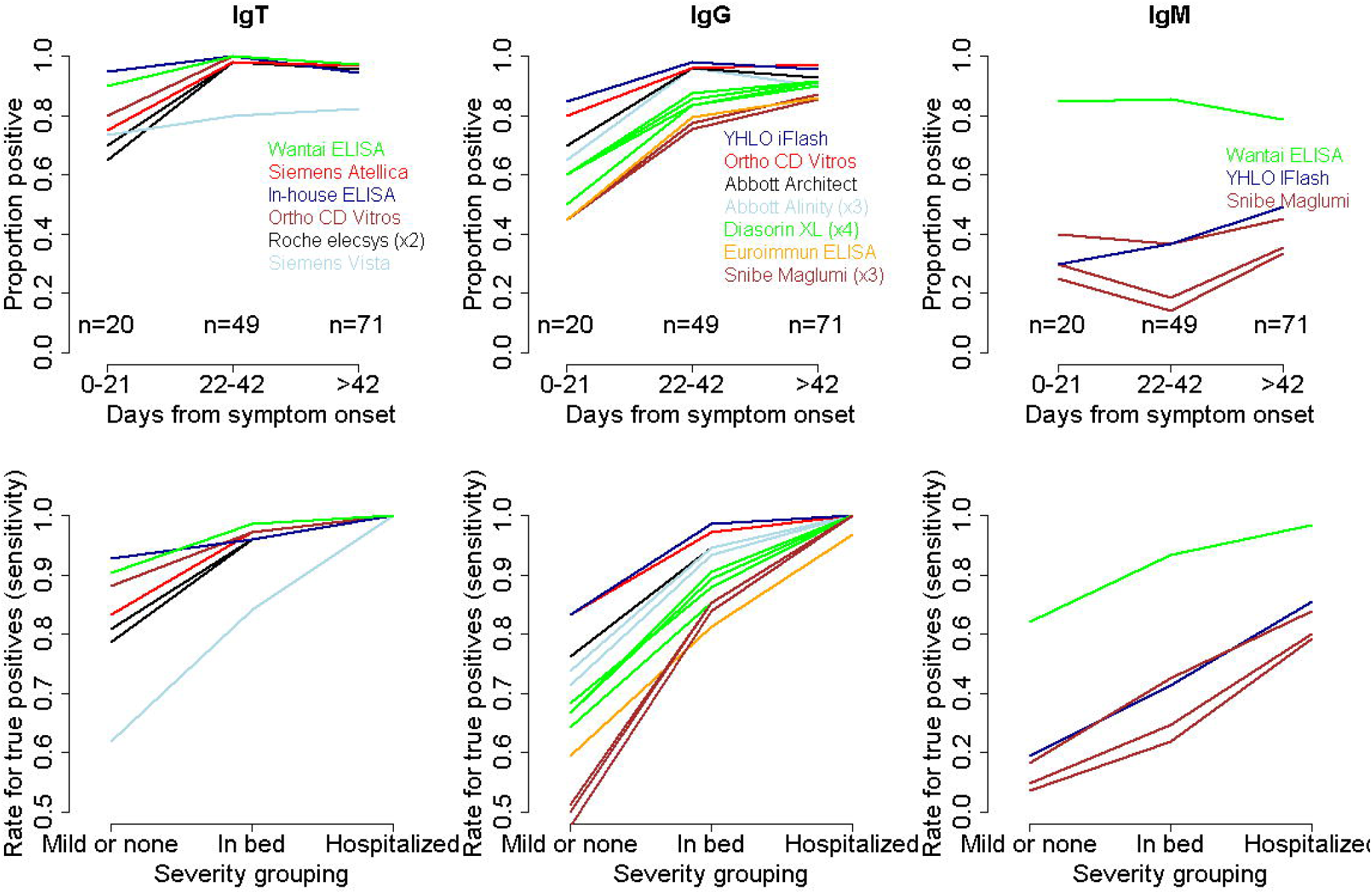
Antibody development for total-Ab (IgT), IgG and IgM as a function of days from symptom onset in three-week periods and severity of symptoms grouped as “mild or none”, “in bed” at home or “hospitalized”. Assays are color coded. Several lines with the same color appear if the same assay was performed in different laboratories as indicated to show the (small) interlaboratory variation.

A total of 4 samples were not detected by any of the assays. Of these, 3 patients had mild clinical symptoms and one patient had moderate symptoms, with the TSO varying between 17-73 days.

## Discussion

The results of this study demonstrated that diagnostic accuracy was highest in the group of the SARS-CoV-2 total-Ab assays compared to the group of the SARS-CoV-2 IgG assays. Except for one outlier - Siemens Vista - all total-Ab assays resulted in acceptable performances for diagnostic use regarding both specificity and sensitivity. The SARS-CoV-2 IgG assays demonstrated a larger variation in sensitivities, and only three assays demonstrated a predefined acceptable sensitivity for diagnostic use, whereas one assay did not meet the defined acceptance criteria for specificity. All total-Ab and IgG assays showed higher seropositive rates in samples from patients with a TSO > 3 weeks, and seropositive rates increased with symptom severity in all assays across all Ig types.

Nominally, the accuracy of the Wantai total-Ab, the in-house CUH/NOVO total-Ab ELISA, and the Ortho CD Vitros total-Ab assays were the best, with optimal sensitivity for diagnostic use according to our criteria. However, the Roche Elecsys total-Ab and the Siemens Atellica total-Ab assays also performed with acceptable sensitivity for diagnostic use, and the confidence intervals for sensitivity and specificity overlapped for the mentioned assays. The poor sensitivity of the Siemens Vista assay was seemingly due to the manufacturer’s setting with a suboptimal assigned cut-off value. For example, adjusting the Siemens Vista assay cut-off from 1000 to 375 increased the sensitivity from 81.0 to 93.9% without changing the 100% specificity, exemplifying the risk of the forced releases of new assays. Adjustment in cut-offs could potentially also improve the performance of other investigated assays. However, for this study, we used assays as specified by the manufacturers of the assays.

Among the SARS-Cov-2 IgG assays, three assays - YHLO iFlash IgG, Ortho CD Vitros IgG, and Abbott Architect IgG - showed acceptable sensitivity performance for diagnostic use. When using a TSO limit of >21 days for the antibody assays, the Abbott Alinity IgG assay also reached an acceptable sensitivity. Regarding specificity, only the Diasorin/LiaisonXL IgG assay did not meet the defined criteria. More than half of the samples in the case panel had a TSO > 6 weeks and were not optimal for the evaluation of the sensitivity of the IgM assays. However, among the IgM assays, the Wantai IgM assay stood out with a relatively high sensitivity and specificity.

In three of the evaluated assays, a recombinant nucleocapsid antigen (rN) is used in the immunoassay, while in eight assays, a recombinant spike antigen (rS) of the RBD is used; two assays did not specify the protein(s) used as the capturing antigen in the assay, and two assays (the YHLO IgG and IgM) use both rN and rS. Our study does not seem to suggest that the chosen antigen (N vs S RBD) affects the assay performance in general. Instead, the differences in performance seem to be associated with overall assay design rather than the choice of antigen. This parallels the observations by Havari, who demonstrated the appearance of neutralizing antibodies against both N and S proteins simultaneously (11).

A potential limitation for the use of immunoassays can be interference due to x-reactivity in individuals with autoimmune disease or acute infections with EBV or CMV. We tested this directly by using samples from patients with these diseases. This did not seem to be an issue in most assays, except for in the Diasorin/LiaisonXL IgG and YHLO/iFlash IgM assays, which showed interference in some samples.

In the US, the FDA (US Food and Drug Administration) requires a minimum sensitivity of 90% and a specificity of 95% for emergency use authorization of serologic anti-SARS-CoV-2 assays (12). In the UK, the Medicines and Healthcare Products Regulatory Agency (MHRA) has defined a Target Product Profile for Enzyme Immunoassays detecting antibodies against SARS-CoV-2, with an acceptable sensitivity and specificity of ≥98% for anti-SARS-CoV-2-immunoassays among patients with a history of SARS-CoV-2 ≥20 days after symptom onset (13). In our study, the sensitivities calculated from the case samples with a known TSO >21 days (N=123) did not reach 98% in any assay evaluated (Table 3a-c). However, we prioritized high diagnostic specificity (≥99%) as our main criterion, since Denmark has a low anti-SARS-CoV-2 seroprevalence (1.7%) (14). For example, in a population with a low anti-SARS-CoV-2 prevalence, an assay specificity of 97.2% (Diasorin/LiaisonXL) would lead to low positive predictive values. Interestingly, many manufacturers use a TSO ≥14 days, in contrast to ≥20 or >21 days, as a cut-off for optimal sensitivity, so there is a need for international consensus on establishing a criterion for TSO to test for optimal sensitivity.

We found 4 cases with no anti-SARS-CoV-2 antibodies in any assay. This finding could be explained by several factors, including an early-stage infection, mild disease, a transient antibody response only, no antibodies produced or produced at nondetectable levels, or false positive NAAT tests. Early stage infection was not the case in these 4 patients, who had a TSO between 17-73 days, whereas the largest variation in sensitivity performance between the anti-SARS-CoV-2 assays was seen in the samples with a TSO ≤ 21 days.

The TSO among the COVID-19 case samples indeed determined the absolute sensitivity values obtained, as the median seroconversion time is reportedly 11 days (interquartile range of 7.3-14.0 days) after onset of symptoms (15-18). Though in this case, with a TSO of 17 to 73 days, this does not seem to be a plausible explanation for the lack of antibodies; instead, a combination of mild COVID-19 symptoms in the nonhospitalized group of case panel members and the collection of blood samples in the late convalescent stage might explain the nondetectable antibodies. As we showed, most anti-SARS-CoV-2 total-Ab and IgG assays had a higher rate of seropositivity among the hospitalized patients compared to the nonhospitalized patients, and previous studies have also shown that anti-SARS-CoV-2 titers correlate with the severity of COVID-19 among hospitalized patients (19, 20) and that there is a time-dependent decline in antibody titers for anti-SARS-CoV-2 Ab in general, including neutralizing antibodies (14, 20).

The data also provides some interesting preliminary observations regarding the antibody response in general. First, most individuals (approximately 97%) seem to develop some sort of antibody response. Second, this response seems to be at a peak in samples taken at approximately 3 weeks after TSO. Third, the response seems to be positively correlated with disease severity. However, while our study compares the ability of antibody assays to detect people who have had NAAT-confirmed COVID-19, it does not compare the ability to detect individuals who are protected against reinfection with SARS-CoV-2. Identification of those who are protected against reinfection, at least for a certain period, would be an important aspect of an assay, but it is too early in the epidemic to make this kind of comparison on a large scale. However, a recent study from GeurtsvanKessel et al. reported cut-off values in the Wantai total-Ab assay indicating detectable levels of neutralizing antibodies. The authors suggest this as a tool for the detection of neutralizing antibodies, though the clinical utility of this remains unclear (21).

Our study has several strengths and limitations. A major strength is that the case panel used for sensitivity across all 16 assays included 150 samples from the same patients, which is one of the largest panels investigated to date. Additionally, and in contrast to most previous evaluations of serological SARS-CoV-2 assays, this case panel was obtained largely from patients who had milder symptoms of COVID-19, evaluating whether the assays could detect SARS-CoV-2 antibodies among the most common type of patient with SARS-CoV-2 infection. This is valuable knowledge in sero-epidemiological investigations. The specificity was evaluated with a significant number of pre-COVID-19 blood donor samples, making this study very solid in terms of clinical accuracy and agreement between the assays investigated. Furthermore, we investigated x-reactivity using samples from individuals and specificity using individuals with autoimmune disease or acute infections with EBV or CMV. For these tests of specificity, we could not use samples from the same individuals across all assays due to the small sample volumes. This could potentially introduce heterogeneity between assay in the specificity data. However, using ≥586 samples from healthy donors for each assay makes this a very large sample and substantial differences in a small homogenous country with same standard operating procedures for the Danish blood banks is unlikely. Even though we tried to assess the risk of interference by examining serum from patients with viral diseases known to be associated with increased levels of assay-interfering antibodies, these could be present in patients with other diseases, e.g., cancers. Thus, we could have underestimated the potential for interference.

In conclusion, this comparative study of 15 commercial and 1 in-house laboratory serological SARS-CoV-2 assays pinpoints differences in accuracy; however, several total-Ab and IgG assays reached predefined criteria for acceptable performance, especially in samples from cases with a TSO over 3 weeks. Additionally, the antibody response seemed to be strongest among patients with more severe disease. Last, it appears as if the use of emergency authorizations has led to release of suboptimal assays in some cases, and simple measures such as optimization of cut-off values could lead to major improvements in performance. Thus, it is possible that optimized versions of some assays may be released in near future.

## Data Availability

The STARD 2015 checklist has been followed

## Acknowledgements

We wish to thank the laboratory technicians in all laboratories who performed the validation of assays in this evaluation for their thorough and effective laboratory performance. Additionally, we wish to thank the head of DEKS (Danish Institute for External Quality Assurance for Laboratories in the health sector), Gitte Henriksen, for distributing the samples from patients with autoimmune diseases.

We wish to thank Charlotte Helgstrand for providing the antigen used in the in-house CUH-NOVO total-Ab ELISA. The plasmid used for synthesizing the SARS CoV2 RBD polypeptide for that assay was made and kindly contributed by the International AIDS Vaccine Initiative (“IAVI”) and provided by the responsible IAVI employee, Joseph Jardine, Scripps Institute, La Jolla, CA, USA.

The development of the CUH-NOVO SARS-CoV-2 Total Ab ELISA was financially supported by grants from the Carlsberg Foundation (CF20-0045) and the Novo Nordisk Foundation (205A0063505).

Some of the assays evaluated were purchased and some were provided for evaluation from the manufacturer without costs. We thank the manufacturers for technical advice and support.

## Ethical approval

The study of samples from patients with former SARS-CoV-2 infection for validation of serological SARS-CoV-2 assays was approved by the Regional Scientific Committee for the Capital Region of Denmark (H-20028627). All blood donors were asked for consent for using archive samples for their use in the validation of new methods and assay investigations as quality control projects.

## References

1. Ghebreyesus T. 2020. WHO Director-General’s opening remarks at the media briefing on COVID-19 - 11 March 2020.

2. Rabi FA, Al Zoubi MS, Kasasbeh GA, Salameh DM, Al-Nasser AD. 2020. SARS-CoV-2 and Coronavirus Disease 2019: What We Know So Far. Pathogens 9.

3. Prompetchara E, Ketloy C, Palaga T. 2020. Immune responses in COVID-19 and potential vaccines: Lessons learned from SARS and MERS epidemic. Asian Pac J Allergy Immunol 38:1–9.

4. Deeks JJ, Dinnes J, Takwoingi Y, Davenport C, Spijker R, Taylor-Phillips S, Adriano A, Beese S, Dretzke J, Ferrante di Ruffano L, Harris IM, Price MJ, Dittrich S, Emperador D, Hooft L, Leeflang MM, Van den Bruel A. 2020. Antibody tests for identification of current and past infection with SARS-CoV-2. Cochrane Database Syst Rev 6:Cd013652.

5. Theel ES, Slev P, Wheeler S, Couturier MR, Wong SJ, Kadkhoda K. 2020. The Role of Antibody Testing for SARS-CoV-2: Is There One? JClinMicrobiol.

6. Whitman JD, Hiatt J, Mowery CT, Shy BR, Yu R, Yamamoto TN, Rathore U, Goldgof GM, Whitty C, Woo JM, Gallman AE, Miller TE, Levine AG, Nguyen DN, Bapat SP, Balcerek J, Bylsma SA, Lyons AM, Li S, Wong AW, Gillis-Buck EM, Steinhart ZB, Lee Y, Apathy R, Lipke MJ, Smith JA, Zheng T, Boothby IC, Isaza E, Chan J, Acenas DD, Lee J, Macrae TA, Kyaw TS, Wu D, Ng DL, Gu W, York VA, Eskandarian HA, Callaway PC, Warrier L, Moreno ME, Levan J, Torres L, Farrington LA, Loudermilk R, Koshal K, Zorn KC, Garcia-Beltran WF, Yang D, et al. 2020. Test performance evaluation of SARS-CoV-2 serological assays. medRxiv.

7. Bastos ML TG, Abidi SK, Campbell J R, Haraoui L, Johnston JC, Lan Z, Law S, MacLean E, Trajman A, Menzies D, Benedetti A, Khan FA. 2020. Diagnostic accuracy of serological tests for covid-19: systematic review and meta-analysis. BMJ 2020;370:m2516 doi:https://www.bmj.com/content/370/bmj.m2516.

8. Ria Lassaunière AF, Zitta B. Harboe, Alex C.Y. Nielsen, Anders Fomsgaard, Karen A. Krogfelt, Charlotte S. Jørgensen. 2020. Evaluation of nine commercial SARS-CoV-2 immunoassays. medRxiv doi:https://www.medrxiv.org/content/10.1101/2020.04.09.20056325v1.

9. Kohmer N, Westhaus S, Ruhl C, Ciesek S, Rabenau HF. 2020. Brief clinical evaluation of six high-throughput SARS-CoV-2 IgG antibody assays. JClinVirol 129:104480.

10. Voldstedlund M, Haarh M, Molbak K, MiBa Board of R. 2014. The Danish Microbiology Database (MiBa) 2010 to 2013. Euro Surveill 19.

11. Haveri A, Smura T, Kuivanen S, Osterlund P, Hepojoki J, Ikonen N, Pitkapaasi M, Blomqvist S, Ronkko E, Kantele A, Strandin T, Kallio-Kokko H, Mannonen L, Lappalainen M, Broas M, Jiang M, Siira L, Salminen M, Puumalainen T, Sane J, Melin M, Vapalahti O, Savolainen-Kopra C. 2020. Serological and molecular findings during SARS-CoV-2 infection: the first case study in Finland, January to February 2020. Euro Surveill 25.

12. Duong YT WC, Justman J. 2020. Antibody testing for coronavirus disease 2019: not ready for prime time. BMJ 2020;370:m2655.

13. https://www.gov.uk/government/organisations/medicines-and-healthcare-products-regulatory-agency. 2020. Target Product Profile EIA antibody tests antibodies to SARS-CoV-2. https://www.gov.uk/government/publications/how-tests-and-testing-kits-for-coronavirus-covid-19-work/target-product-profile-enzyme-immunoassay-eia-antibody-tests-to-help-determine-if-people-have-antibodies-to-sars-cov-2. Accessed

14. Erikstrup C, Hother CE, Pedersen OBV, Molbak K, Skov RL, Holm DK, Saekmose SG, Nilsson AC, Brooks PT, Boldsen JK, Mikkelsen C, Gybel-Brask M, Sorensen E, Dinh KM, Mikkelsen S, Moller BK, Haunstrup T, Harritshoj L, Jensen BA, Hjalgrim H, Lillevang ST, Ullum H. 2020. Estimation of SARS-CoV-2 infection fatality rate by real-time antibody screening of blood donors. ClinInfectDis.

15. Long QX, Liu BZ, Deng HJ, Wu GC, Deng K, Chen YK, Liao P, Qiu JF, Lin Y, Cai XF, Wang DQ, Hu Y, Ren JH, Tang N, Xu YY, Yu LH, Mo Z, Gong F, Zhang XL, Tian WG, Hu L, Zhang XX, Xiang JL, D. HX, Liu HW, Lang CH, Luo XH, Wu SB, Cui XP, Zhou Z, Zhu MM, Wang J, Xue CJ, Li XF, Wang L, Li ZJ, Wang K, Niu CC, Yang QJ, Tang XJ, Zhang Y, Liu XM, Li JJ, Zhang DC, Zhang F, Liu P, Yuan J, Li Q, Hu JL, Chen J, et al. 2020. Antibody responses to SARS-CoV-2 in patients with COVID-19. Nat Med 26:845–848.

16. Zhao J, Yuan Q, Wang H, Liu W, Liao X, Su Y, Wang X, Yuan J, Li T, Li J, Qian S, Hong C, Wang F, Liu Y, Wang Z, He Q, Li Z, He B, Zhang T, Fu Y, Ge S, Liu L, Zhang J, Xia N, Zhang Z. 2020. Antibody responses to SARS-CoV-2 in patients of novel coronavirus disease 2019. ClinInfectDis.

17. Lou B, Li TD, Zheng SF, Su YY, Li ZY, Liu W, Yu F, Ge SX, Zou QD, Yuan Q, Lin S, Hong CM, Yao XY, Zhang XJ, Wu DH, Zhou GL, Hou WH, Li TT, Zhang YL, Zhang SY, Fan J, Zhang J, Xia NS, Chen Y. 2020. Serology characteristics of SARS-CoV-2 infection since exposure and post symptom onset. Eur Respir J doi:10.1183/13993003.00763-2020.

18. Huang AT, Garcia-Carreras B, Hitchings MDT, Yang B, Katzelnick LC, Rattigan SM, Borgert BA, Moreno CA, Solomon BD, Rodriguez-Barraquer I, Lessler J, Salje H, Burke D, Wesolowski A, Cummings DAT. 2020. A systematic review of antibody mediated immunity to coronaviruses: antibody kinetics, correlates of protection, and association of antibody responses with severity of disease. medRxiv doi:10.1101/2020.04.14.20065771.

19. Qu J, Wu C, Li X, Zhang G, Jiang Z, Li X, Zhu Q, Liu L. 2020. Profile of IgG and IgM antibodies against severe acute respiratory syndrome coronavirus 2 (SARS-CoV-2). Clin Infect Dis doi:10.1093/cid/ciaa489.

20. Seow J GC, Merrick B, Acors S, Steel KJA, Hemmings O, O’Bryne A, Kouphou N, Pickering S, Galao R, Betancor G, Wilson HD, Signell AW, Winstone H, Kerridge C, Temperton N, Snell L, Bisnauthsing K, Moore A, Green A, Martinez L, Stokes B, Honey J, Izquierdo-Barras A, Arbane G, Patel A, OConnell L, O’Hara G, MacMahon E, Douthwaite S, Nebbia S, Batra R, Martinez-Nunez R, Edgeworth JD, Neil SJD, Malim M, Doores K. 2020. Longitudinal evaluation and decline of antibody responses in SARS-CoV-2 infection. medRxiv doi:https://doi.org/10.1101/2020.07.09.20148429.

21. GeurtsvanKessel CH, Okba NMA, Igloi Z, Bogers S, Embregts CWE, Laksono BM, Leijten L, Rokx C, Rijnders B, Rahamat-Langendoen J, van den Akker Jpc, van Kampen Jja, van der Eijk AA, van Binnendijk RS, Haagmans B, Koopmans M. 2020. An evaluation of COVID-19 serological assays informs future diagnostics and exposure assessment. NatCommun 11:3436.

